# Sagittal balance of the spine - a geometric force-line model

**DOI:** 10.1101/2025.01.24.25321026

**Authors:** Kai Song, Yu Wang, Qiang Yang, Cao Yang, Bing Wang, Fangcai Li, Zezhang Zhu, Weishi Li, Jianguo Zhang, Zheng Wang

**Affiliations:** Department of Orthopaedics, Chinese People’s Liberation Army General Hospital, Beijing, PR China; Department of Orthopedics, Peking University First Hospital, Beijing, PR China; Department of Spine Surgery, Tianjin Hospital, Tianjin, PR China; Department of Orthopaedics, Union Hospital, Tongji Medical College, Huazhong University of Science and Technology, Wuhan, Hubei, PR China; Department of Spine Surgery, Second Xiangya Hospital of Central South University, Changsha, Hunan, PR China; Department of Orthopedics Surgery, Second Affiliated Hospital, School of Medicine, Zhejiang University, Hangzhou, Zhejiang, PR China; Department of Spine Surgery, The Affiliated Drum Tower Hospital of Nanjing University Medical School, Nanjing, Jiangsu, PR China; Department of Orthopaedics, Peking University Third Hospital, Beijing, PR China; Department of Orthopedics, Peking Union Medical College Hospital, Beijing, PR China

**Author notes:** Kai Song* ✉. Wang Zheng ✉.

**Keywords:** Sagittal balance, Spinal-pelvic alignments, Lumbosacral lordosis

## Abstract

**Objective:** To introduce a new geometric force-line model to assess the sagittal spinal-pelvic alignments and statistically analyze the main parameters of the model, offering a novel perspective for understanding the sagittal balance.

**Methods:** A total of 388 healthy adults aged 18 - 35 years were recruited. To designate the mid-point of the anterior edge of the S2 vertebral body as the imaging marker point of the sagittal sacroiliac joint (SIJ). Parameters were measured included lines’ inclinations and angles related to T1, S2, HA (hip axis) and Co1(the first coccygeal).

**Results:** The S2-T1 inclination angle was 89.5° ± 2.8°, close to perpendicular (90°), and it means the spinal axis (S2-T1) of the population is approximately perpendicular to the ground. In addition, spinal axis (S2-T1) crossed T11-L1 segments, which means spinal S curve is basically distributed on both sides of the axis. Angles ∠Co1-S2-HA, ∠HA-Co1-S2, and ∠S2-HA-Co1 were approximate 60°, forming a horizontal stable equilateral triangle. The anterior edge of S2 was nearly perpendicular to the pelvic force line (∠S2, HA-S2 ≈ 90°).

**Conclusion:** The geometric performance of the proposed model is concise, clear, and logical. It provides a new perspective on the sagittal balance. This framework might help us better understand spinal-pelvic alignments and its clinical significance.

## Introduction

A profound comprehension of the sagittal alignment of the human spine and pelvis holds pivotal significance for the diagnosis and treatment of spinal disorders **[1, 2]**. The sagittal spinal parameters measured with the Cobb method, including thoracic kyphosis (TK) and lumbar lordosis (LL), along with sagittal pelvic parameters such as pelvic incidence (PI), sacral slope (SS), and pelvic tilt (PT), and the sagittal vertical axis (SVA), have emerged as the standard metrics for assessing the sagittal alignment of the spine and pelvis **[3, 4]**. Furthermore, theoretical frameworks for sagittal balance reconstruction, such as the Roussouly classification, Schwab - SRS classification, and Gap scoring system, all of which are derived from these parameters, have been extensively implemented in clinical practice by spine surgeons **[5 – 7]**. Nevertheless, postoperative complications and unforeseen circumstances associated with sagittal alignment remain prevalent **[8, 9]**. Consequently, there is an urgent necessity to explore a more rational theoretical system for the sagittal balance of the spine and pelvis.

The sagittal “S” curve of the spine has been a crucial factor in the historical transition of human evolution from a quadrupedal to a bipedal upright posture **[10 – 12]**. Song et al. posited that the distal end of the “S” curve terminates at S2, thus suggesting that its defined range should be adjusted from the previous “LL + TK” to “lumbosacral lordosis (LSL) + TK” **[13, 14]**. Based on this, the straight line connecting S2 and T1 is likely to approximate the spinal axis (force line).

In addition, the evolution of the pelvic structure, including the hip joint and sacroiliac joint, has also played a significant role in the development of human upright posture **[10 – 12, 15]**. The hip axis (HA) serves as the imaging connection point between the pelvis and the lower limbs, while the sacroiliac joint (SIJ) functions as the connection point between the spine and the pelvis. However, it is challenging to identify the imaging projection of the central point of the SIJ in the lateral view. Drawing on previous research **[13, 14]** and logical deductions, the author designates the mid - point of the anterior edge of the S2 vertebral body as the imaging marker point of the sagittal SIJ.

Based on these three points (T1, SIJ or S2, HA) and two lines (S2-T1, HA-S2), the author proposes and establishes a sagittal spinal-pelvic geometric force-line model **(Figure 1)**. This study aims to measure and statistically analyze the main parameters of this model, offering a novel perspective for understanding the sagittal balance of the spine and pelvis.

**Figure 1.**
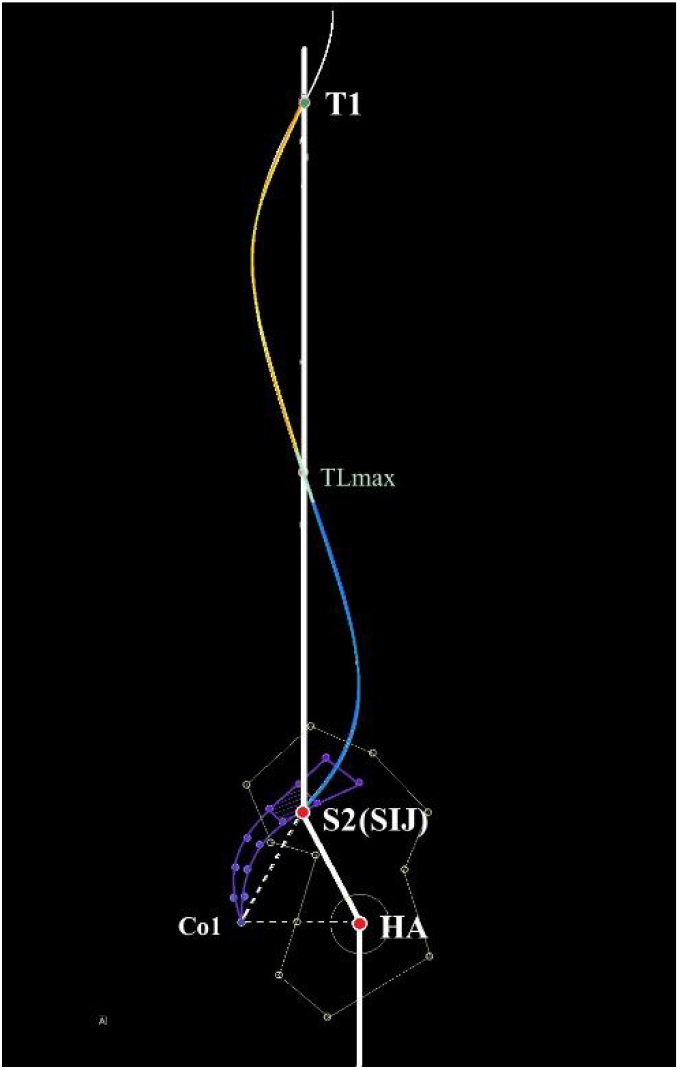
Schematic Representation of the Sagittal Spinal - Pelvic Geometric Force - Line Model T1: The mid - point of the superior end - plate of the first thoracic vertebra. **S2:** The mid - point of the anterior margin of the second sacral vertebra. It is employed as the radiographic marker for the sacroiliac joint (**SIJ**). Given the difficulty in precisely identifying the SIJ’s central point in lateral imaging, this mid-point of the S2 anterior margin provides a practical and consistent landmark for analysis. **HA**: Hip axis, which is defined as the mid-point of the line connecting the centers of the bilateral femoral heads. **Co1**: The distal apex of the first coccygeal vertebra. **TL max**: The maximum horizontal angle of posterior inclination of the thoracic and lumbar vertebral segments.

## Methods

A total of 388 healthy adults aged 18 - 35 years were included in this study, and their full-length lateral spinal rays in free-standing position were collected. Based on the maximum thoracic and lumbar inclination angle (TL max, the angle between the line connecting the mid-points of the upper and lower end-plates of the most inclined vertebral body and the right-hand horizontal line), the subjects were grouped as follows: Group A < 106.5°; 106.5° ≤ Group B < 114.5°; Group C ≥ 114.5°.

The following parameters were measured and statistically analyzed: TL max, lumbosacral lordosis (LSL, measured as the Ferguson angle between L1 and S2), thoracic kyphosis (TK, measured as the Ferguson angle between T1 and T12); the inclination angle (the inclination angle is defined as the angle with the right-hand horizontal line) of S2-T1, Co1-HA, Co1-S2, HA-S2 and S2 (anterior edge of S2); ∠Co1-S2-HA, ∠HA-Co1-S2, ∠S2-HA-Co1; and ∠S2-T1, Co1-HA, ∠Co1-S2, S2-T1, ∠HA-S2, S2-T1, ∠S2, S2-HA; ∠S2-L1L, L1L-T1 (L1L is defined as the mid-point of the lower end-plate of L1), ∠S2-L1, L1-T1, ∠S2-L1U, L1U-T1 (L1U is defined as the mid-point of the upper end-plate of L1), ∠S2-T12L, T12L-T1, ∠S2-T12, T12-T1, ∠S2-T11/12, T11/12-T1 ( T11/12 is defined as the mid-point of the T11-T12 intervertebral disc), ∠S2-T11, T11-T1, ∠S2-T11U, T11U-T1.

The mean value, standard deviation, and extreme values of each parameter were calculated, and a chi-square test was performed for comparative analysis among the groups.

## Results

Based on TL max grouping: The TL max of all subjects was 110.5±5.5° (range 94.4-132.0°). Group A consisted of 91 cases with a TL max of 104.0±2.3° (range 94.4-106.5°); Group B had 216 cases with a TL max of 110.2±2.1° (range 106.6-114.5°); Group C included 81 cases with a TL max of 118.6±3.8° (range 114.7-132.0°). The S2-T1 inclination angle for all subjects was 89.5±2.8° (range 81.5-97.9°), with Group A at 87.6±2.6° (range 81.5-93.5°), Group B at 89.5±2.4° (range 81.6-96.4°), and Group C at 91.8±2.6° (range 86.3-97.9°). The HA-S2 inclination angle for all subjects was 121.8±8.0° (range 97.1-144.3°), with Group A at 122.4±7.8° (range 102.1-144.3°), Group B at 122.3±7.7° (range 98.6-141.1°), and Group C at 119.8±8.8° (range 97.1-135.7°). There were no statistically significant differences among the three groups when p<0.01. The angles ∠Co1-S2-HA, ∠ HA-Co1-S2, and ∠S2-HA-Co1 were approximately equal to 60°, with no significant statistical differences among Groups A, B, and C. Detailed results are presented in **Table 1** and **Figure 2**.

**Table 1.**
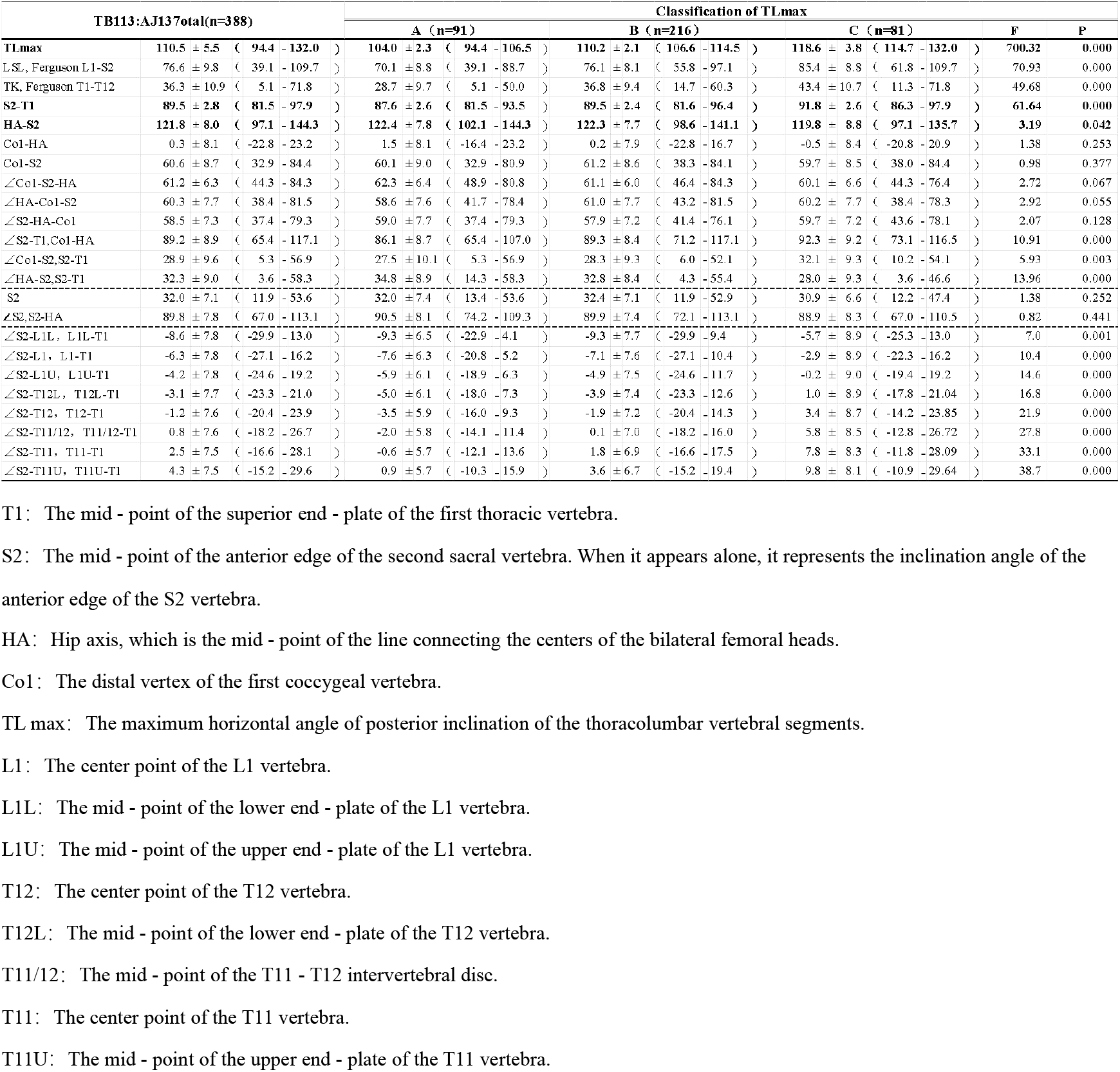
Statistical Description of the Geometric Relationships of Force - Line.

| TB113:AJ137total(n=388) |  | Classification of TLmax |  |  |  |  |
| --- | --- | --- | --- | --- | --- | --- |
|  |  | A (n=91) | B (n=216) | C (n=81) | F | P |
| TLmax | 110.5 ± 5.5 ( 94.4 - 132.0 ) | 104.0 ± 2.3 ( 94.4 - 106.5 ) | 110.2 ± 2.1 ( 106.6 - 114.5 ) | 118.6 ± 3.8 ( 114.7 - 132.0 ) | 700.32 | 0.000 |
| L-SL, Ferguson L1-S2 | 76.6 ± 9.8 ( 39.1 - 109.7 ) | 70.1 ± 8.8 ( 39.1 - 88.7 ) | 76.1 ± 8.1 ( 55.8 - 97.1 ) | 85.4 ± 8.8 ( 61.8 - 109.7 ) | 70.93 | 0.000 |
| TK, Ferguson T1-T12 | 36.3 ± 10.9 ( 5.1 - 71.8 ) | 28.7 ± 9.7 ( 5.1 - 50.0 ) | 36.8 ± 9.4 ( 14.7 - 60.3 ) | 43.4 ± 10.7 ( 11.3 - 71.8 ) | 49.68 | 0.000 |
| S2-T1 | 89.5 ± 2.8 ( 81.5 - 97.9 ) | 87.6 ± 2.6 ( 81.5 - 93.5 ) | 89.5 ± 2.4 ( 81.6 - 96.4 ) | 91.8 ± 2.6 ( 86.3 - 97.9 ) | 61.64 | 0.000 |
| HA-S2 | 121.8 ± 8.0 ( 97.1 - 144.3 ) | 122.4 ± 7.8 ( 102.1 - 144.3 ) | 122.3 ± 7.7 ( 98.6 - 141.1 ) | 119.8 ± 8.8 ( 97.1 - 135.7 ) | 3.19 | 0.042 |
| Co1-HA | 0.3 ± 8.1 ( -22.8 - 23.2 ) | 1.5 ± 8.1 ( -16.4 - 23.2 ) | 0.2 ± 7.9 ( -22.8 - 16.7 ) | -0.5 ± 8.4 ( -20.8 - 20.9 ) | 1.38 | 0.253 |
| Co1-S2 | 60.6 ± 8.7 ( 32.9 - 84.4 ) | 60.1 ± 9.0 ( 32.9 - 80.9 ) | 61.2 ± 8.6 ( 38.3 - 84.1 ) | 59.7 ± 8.5 ( 38.0 - 84.4 ) | 0.98 | 0.377 |
| ∠Co1-S2-HA | 61.2 ± 6.3 ( 44.3 - 84.3 ) | 62.3 ± 6.4 ( 48.9 - 80.8 ) | 61.1 ± 6.0 ( 46.4 - 84.3 ) | 60.1 ± 6.6 ( 44.3 - 76.4 ) | 2.72 | 0.067 |
| ∠HA-Co1-S2 | 60.3 ± 7.7 ( 38.4 - 81.5 ) | 58.6 ± 7.6 ( 41.7 - 78.4 ) | 61.0 ± 7.7 ( 43.2 - 81.5 ) | 60.2 ± 7.7 ( 38.4 - 78.3 ) | 2.92 | 0.055 |
| ∠S2-HA-Co1 | 58.5 ± 7.3 ( 37.4 - 79.3 ) | 59.0 ± 7.7 ( 37.4 - 79.3 ) | 57.9 ± 7.2 ( 41.4 - 76.1 ) | 59.7 ± 7.2 ( 43.6 - 78.1 ) | 2.07 | 0.128 |
| ∠S2-T1, Co1-HA | 89.2 ± 8.9 ( 65.4 - 117.1 ) | 86.1 ± 8.7 ( 65.4 - 107.0 ) | 89.3 ± 8.4 ( 71.2 - 117.1 ) | 92.3 ± 9.2 ( 73.1 - 116.5 ) | 10.91 | 0.000 |
| ∠Co1-S2, S2-T1 | 28.9 ± 9.6 ( 5.3 - 56.9 ) | 27.5 ± 10.1 ( 5.3 - 56.9 ) | 28.3 ± 9.3 ( 6.0 - 52.1 ) | 32.1 ± 9.3 ( 10.2 - 54.1 ) | 5.93 | 0.003 |
| ∠HA-S2, S2-T1 | 32.3 ± 9.0 ( 3.6 - 58.3 ) | 34.8 ± 8.9 ( 14.3 - 58.3 ) | 32.8 ± 8.4 ( 4.3 - 55.4 ) | 28.0 ± 9.3 ( 3.6 - 46.6 ) | 13.96 | 0.000 |
| S2 | 32.0 ± 7.1 ( 11.9 - 53.6 ) | 32.0 ± 7.4 ( 13.4 - 53.6 ) | 32.4 ± 7.1 ( 11.9 - 52.9 ) | 30.9 ± 6.6 ( 12.2 - 47.4 ) | 1.38 | 0.252 |
| ∠S2, S2-HA | 89.8 ± 7.8 ( 67.0 - 113.1 ) | 90.5 ± 8.1 ( 74.2 - 109.3 ) | 89.9 ± 7.4 ( 72.1 - 113.1 ) | 88.9 ± 8.3 ( 67.0 - 110.5 ) | 0.82 | 0.441 |
| ∠S2-L1L, L1L-T1 | -8.6 ± 7.8 ( -29.9 - 13.0 ) | -9.3 ± 6.5 ( -22.9 - 4.1 ) | -9.3 ± 7.7 ( -29.9 - 9.4 ) | -5.7 ± 8.9 ( -25.3 - 13.0 ) | 7.0 | 0.001 |
| ∠S2-L1, L1-T1 | -6.3 ± 7.8 ( -27.1 - 16.2 ) | -7.6 ± 6.3 ( -20.8 - 5.2 ) | -7.1 ± 7.6 ( -27.1 - 10.4 ) | -2.9 ± 8.9 ( -22.3 - 16.2 ) | 10.4 | 0.000 |
| ∠S2-L1U, L1U-T1 | -4.2 ± 7.8 ( -24.6 - 19.2 ) | -5.9 ± 6.1 ( -18.9 - 6.3 ) | -4.9 ± 7.5 ( -24.6 - 11.7 ) | -0.2 ± 9.0 ( -19.4 - 19.2 ) | 14.6 | 0.000 |
| ∠S2-T12L, T12L-T1 | -3.1 ± 7.7 ( -23.3 - 21.0 ) | -5.0 ± 6.1 ( -18.0 - 7.3 ) | -3.9 ± 7.4 ( -23.3 - 12.6 ) | 1.0 ± 8.9 ( -17.8 - 21.04 ) | 16.8 | 0.000 |
| ∠S2-T12, T12-T1 | -1.2 ± 7.6 ( -20.4 - 23.9 ) | -3.5 ± 5.9 ( -16.0 - 9.3 ) | -1.9 ± 7.2 ( -20.4 - 14.3 ) | 3.4 ± 8.7 ( -14.2 - 23.85 ) | 21.9 | 0.000 |
| ∠S2-T11/12, T11/12-T1 | 0.8 ± 7.6 ( -18.2 - 26.7 ) | -2.0 ± 5.8 ( -14.1 - 11.4 ) | 0.1 ± 7.0 ( -18.2 - 16.0 ) | 5.8 ± 8.5 ( -12.8 - 26.72 ) | 27.8 | 0.000 |
| ∠S2-T11, T11-T1 | 2.5 ± 7.5 ( -16.6 - 28.1 ) | -0.6 ± 5.7 ( -12.1 - 13.6 ) | 1.8 ± 6.9 ( -16.6 - 17.5 ) | 7.8 ± 8.3 ( -11.8 - 28.09 ) | 33.1 | 0.000 |
| ∠S2-T11U, T11U-T1 | 4.3 ± 7.5 ( -15.2 - 29.6 ) | 0.9 ± 5.7 ( -10.3 - 15.9 ) | 3.6 ± 6.7 ( -15.2 - 19.4 ) | 9.8 ± 8.1 ( -10.9 - 29.64 ) | 38.7 | 0.000 |

T1: The mid - point of the superior end - plate of the first thoracic vertebra.
S2: The mid - point of the anterior edge of the second sacral vertebra. When it appears alone, it represents the inclination angle of the anterior edge of the S2 vertebra.
HA: Hip axis, which is the mid - point of the line connecting the centers of the bilateral femoral heads.
TL max: The maximum horizontal angle of posterior inclination of the thoracolumbar vertebral segments.
L1: The center point of the L1 vertebra.
L1L: The mid - point of the lower end - plate of the L1 vertebra.
L1U: The mid - point of the upper end - plate of the L1 vertebra.
T12: The center point of the T12 vertebra.
T12L: The mid - point of the lower end - plate of the T12 vertebra.
T11/12: The mid - point of the T11 - T12 intervertebral disc.
T11: The center point of the T11 vertebra.
T11U: The mid - point of the upper end - plate of the T11 vertebra.

**Figure 2.**
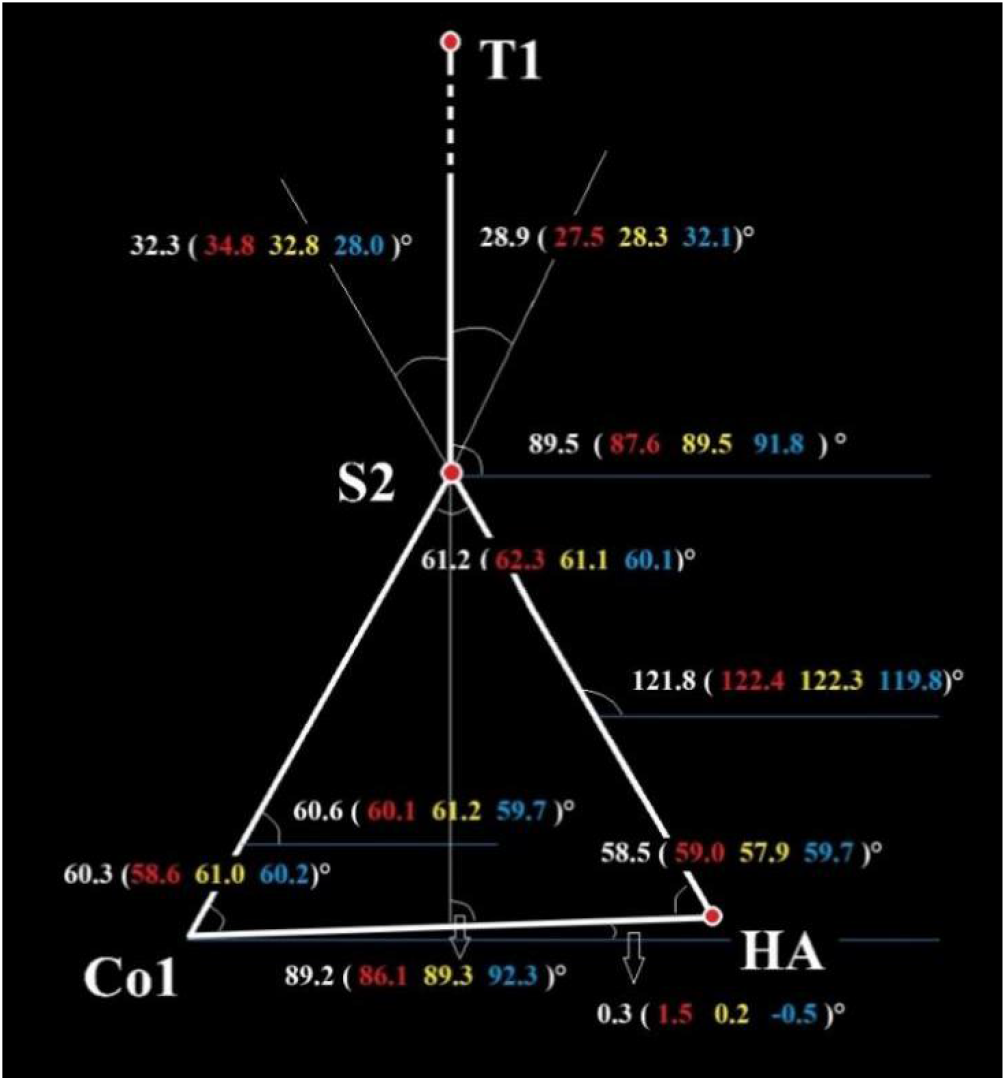
Geometric Relationship Diagram of the Spinal-Pelvic Force-Line Model. The white numbers represent the overall average, the red ones represent the Group A, the yellow ones represent the Group B, and the blue ones represent the Group C. T1: The mid-point of the superior end - plate of the first thoracic vertebra. S2: The mid-point of the anterior edge of the second sacral vertebra. HA: Hip axis, the mid-point of the line connecting the centers of the bilateral femoral heads. Co1: The distal vertex of the first coccygeal vertebra.

In addition, in Groups A, B, and C, the angle ∠S2, HA-S2 was approximately 90° **(Table 1)**, indicating that the anterior edge of the S2 vertebra was essentially perpendicular to the pelvic force line. According to the data in **Table 1**, under mean conditions, in Groups A, B, and C, the spinal axis S2-T1 passed through the thoracolumbar junction (T11-L1) of the spinal sequence.

## Discussion

The structural changes of the spine and pelvis have played an important role in the evolution of humans from quadrupedal to bipedal upright posture **[10 – 12, 15]**.

The “S” shape of the spine provides a structural basis for the mechanical stability and good functional mobility of the upright spine. It was previously believed that the sacrum had a overall kyphotic morphology, and thus the lower end vertebra of the lumbar lordosis (LL) terminated at the superior end - plate of S1. Therefore, the concept range of the “S” curve was considered to be “LL + TK” **[15 - 17]**. However, Song et al. pointed out that the proximal sacrum (S1, S2) also continues the lumbar lordosis and still belongs to the lordotic sequence of the S curve. S2 can be regarded as the end vertebra of the lumbosacral lordosis (LSL) and can be approximately considered as the stress center of the sacroiliac joint. Therefore, its concept range should be defined as “LSL + TK”. Based on the above and using the Ferguson measurement method, their research also showed that LSL is not affected by the size of the pelvic incidence (PI) like LL. Meanwhile, TK is also not affected by PI. Furthermore, PI does not affect the overall curvature of the human S curve. If it is necessary to further distinguish and study the overall curvature of the S curve, the maximum thoracolumbar inclination angle (TL max) can be used as an effective parameter **[14]**. Therefore, in this study, the sagittal spinal sequence was divided into three groups using TL max, and there were significant differences in the overall S curve curvature among the groups. The results of this study showed that the spinal axis (S2-T1) of the population is approximately perpendicular to the ground. Although there are differences in the spinal axes among the groups, they are generally perpendicular to the ground, and the S curves are basically evenly distributed on both sides of the axis, similar to adolescent idiopathic scoliosis (AIS) with double curves in good coronal balance. This is one of the manifestations of a good spinal sequence and is also the operating principle of spinal self-compensation, which will help us understand the state of the sagittal sequence of the spine itself and the compensation mechanism.

The evolution of the pelvic structure has also played an important role in the process of human upright posture **[10 – 12, 15]**. Considering the force-line hinge structure, the hip axis (HA) can be regarded as the linkage point between the pelvis and the lower limbs in imaging. The sacroiliac joint (SIJ) can serve as the linkage point between the pelvis and the spine. Since the imaging projection of the center of the SIJ is difficult to find and identify, based on previous studies and inferences, we used the projection of the mid-point of the anterior edge of S2 in the lateral view as the imaging linkage point of the sacroiliac joint **[13, 14]**. The results of this study confirmed that ΔHA-S2-Co1 is approximately a horizontal and stable equilateral triangle. We can understand HA-S2 as the main structural body of the weight-bearing force line, the S2-Co1 side formed by the curved sacrococcyx is similar to a lever that moves the spine, and Co1-HA can be considered as the tension band that pulls and stabilizes the lever. Even for spines with different curvatures, HA-S2-Co1 is still close to an equilateral triangle. This is one of the manifestations of a normal pelvic sequence, which will help us understand the state of the sagittal sequence and compensation mechanism of the hip joint and sacroiliac joint.

When evaluating the sagittal sequence and balance of an individual’s spine and pelvis, the geometric relationships constructed by three points (T1, S2, HA) and two lines (S2-T1, HA-S2) will be extremely important. The key point and difficulty of this model lie in the uncertainty of the SIJ. In this study, the mid-point of the anterior edge of the S2 vertebral body was used as a general substitute for the SIJ, which is the group performance of the study subjects. For example, the performance of Caucasian and African-American populations may be located at the upper part of S2 or even at the S1/S2 junction. In actual clinical practice, there are differences in the development of the sacrum and sacroiliac joints among individuals. Therefore, the marking points of the sacroiliac joints vary from person to person. The imaging mid-point of the sacroiliac joint may actually fall on the upper part of S2, at the S1/S2 junction, on S1, or on the lower part of S2, or even on S3, or it may be behind or in front of the sacrum. Under normal circumstances, the stress center of the sacrum usually at the inflection point of sagittal alignment and is perpendicular to the pelvic force line and has the shortest distance, as shown in the result of this study where ∠S2, HA - S2 ≈ 90°. A more personalized method of identification is to locate the intersection point of the arcuate line and the sacrum, as well as the inferior posterior iliac spine. The mid-point between these two can be used as an imaging marker for the SIJ in sagittal plane **(Figure 3)**. T1 is usually relatively stable as the upper end vertebra of the spinal axis, but it can also move towards the thoracic and cervical vertebrae when the sagittal spinal sequence is significantly abnormal. HA is absolutely fixed.

**Figure 3.**
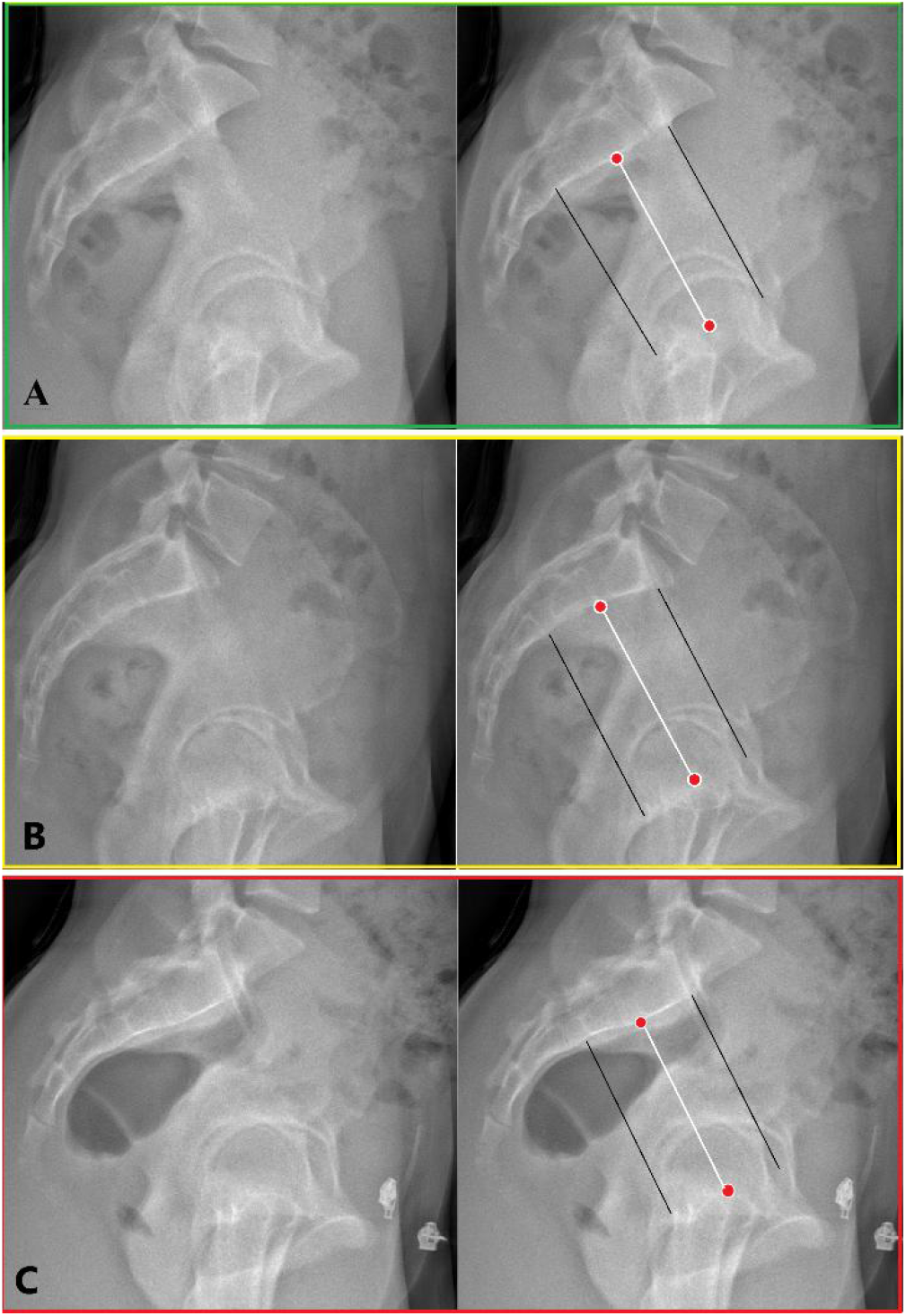
Methods for finding personalized pelvic force line. The two black lines are respectively: the line connecting the intersection of the anterior edge of the sacrum and the arcuate line with the superior edge of the acetabulum; the line connecting the inferior posterior iliac spine with the inferior edge of the acetabulum. These two black lines represent the pelvic force columns, and the white line is their geometric substitute line. A. The SIJ (Sacroiliac Joint) marker is located at the mid - point of the anterior edge of S2. B. The SIJ marker is located at the cephalic end of the anterior edge of S2. C. The SIJ marker is close to the lower end of the anterior edge of S2.

Shortcomings of this paper: The main purpose of this paper is to introduce a new spinal - pelvic force - line model, and its specific details, differences among groups, and clinical applications have not been elaborated. In addition, the cervical force line does not simply continue the spinal axis, and it is closely related to the head, inner ear, and visual field. For the lower limb force line connected to HA, it is usually approximately perpendicular to the ground but is closely related to the functions of the hip, knee, and ankle joints. To avoid excessive length and ensure readability, the head, neck, and lower limb force lines are not included in this model. The author will introduce them separately in subsequent articles.

## Data Availability

All data produced in the present work are contained in the manuscript

